# Core group dynamics of gonorrhea infection in a North Carolina county from 2018 to 2023

**DOI:** 10.1101/2025.04.14.25325476

**Authors:** Brinkley Raynor Bellotti, Jennifer Wenner, Cindy Toler, Vonda Pabon, Michael E. DeWitt, Candice J. McNeil

**Author notes:** **Corresponding author:** Brinkley Raynor Bellotti, Wake Forest University School of Medicine, Medical Center Blvd, Winston-Salem, NC 27157. These authors contributed equally (Michael E. DeWitt, Candice J. McNeil).

## Abstract

**Background:** Core groups, characterized by high levels of sexual activity and disease prevalence, are critical to the transmission dynamics of sexually transmitted infections (STIs) like gonorrhea. Understanding core group dynamics is essential for developing targeted interventions and understanding their potential effect on outcomes for gonorrhea. We assessed core group composition among gonorrhea cases in Guilford County, North Carolina, where gonorrhea rates are elevated, and antimicrobial resistance is a growing concern.

**Methods:** We used *Neisseria gonorrhoeae* surveillance data collected in both public health clinics and emergency departments in Guilford County to characterize core groups. Data including demographics, behavioral risks, and longitudinal gonorrhea test results, were reviewed for specimens submitted for gonorrhea testing. Statistical analyses, including descriptive statistics and univariate logistic regression, were performed to assess factors influencing core group membership within three subpopulations: teens, adult men who have sex with men (MSM), and adult heterosexuals.

**Results:** The study included 25,520 patients, with 1,718 (6.7%) classified as part of the core group. The core group accounted for 68% of all gonorrhea cases, with subgroups such as teen, adult MSM, and adult heterosexual core groups contributing disproportionately to their respective gonorrhea cases (66%, 79%, and 67%, respectively). Demographic and behavioral risk factors are explored and factors increasing odds ratios of core group inclusion are identified among the general population as well as three subpopulations: teens, adult MSM, and adult heterosexual core groups. Notable risk factors for gonorrhea core group inclusion across all subgroups include increased number of sexual partners, drug use, and HIV status. Notably, there is variation in some risk factors among subpopulations including race/ethnicity and sex while traveling internationally.

**Discussion:** Among other factors, variations in public health policies and population level behavioral norms change the way gonorrhea spreads through a population. We report gonorrhea core group composition for a region with high gonorrhea rates. In particular, we discuss the role of teenagers in gonorrhea transmission dynamics, a typically under reported group.

Understanding population specific dynamics is essential for formulating effective and targeted interventions.

## Background

Core groups play a critical role in the transmission dynamics of sexually transmitted infections (STIs), as they are typically characterized by high levels of sexual activity and disease prevalence^1,2^. Understanding the composition of these groups is essential for effective disease control strategies, as factors such as recruitment, preventive behaviors, and the proportion of infected individuals can significantly influence the spread and management of STIs^1,2^. Studies have found that targeted interventions within the in-group can have an outsized effect on disease transmission rather than broad, population level interventions^3–5^. Quantification and description of individuals who compose the core group can lead to more fine-grained, effective interventions to reduce the burden of disease. However, sexual networks are often difficult to quantify, and behaviors and practices vary with the different cultures and practices within the geography^6,7^. As such, understanding of the in-group dynamic within different groups and geographies is not well described for gonorrhea.

Gonorrhea has unique features important to the core group composition: substantial asymptomatic transmission, no measurable immunity after infection, long time course before spontaneous recovery without treatment.^2,8^ For these reasons, the gonorrhea core group is often defined as people with repeat infections, as they are the ones most likely to be infectious over long time periods^8,9^. Core groups have been linked to the expansion of antimicrobial resistant strains of *Neisseria gonorrhoeae*, a growing public health threat^8,10^. In the United States, the Southeast has the highest rates of gonorrhea with state rates ranging from 186 cases per 100,000 (Tennessee) to 288 cases per 100,000 (Louisiana) according to the last STI report published using 2023 data^11^. Given the high morbidity burden in the US Southeast and increasing threat of antimicrobial resistance to frontline treatments, it is vital to understand gonorrhea core group dynamics to formulate targeted interventions.

Across the United States, gonorrhea is most prevalent in men where the majority of cases are symptomatic^12^, and most prevalent in the 20-24 age group according to the 2023 United States Center for Disease Control and Prevention (CDC)^11^. In the 2023 STI report, North Carolina was among the states with the highest rates of gonorrhea^11^. Guilford County, North Carolina was selected as one of the eight jurisdictions in the Centers for Disease Control and Prevention’s initiative “Strengthening the US Response to Resistant Gonorrhea (SURRG).^10^ Guilford county is the third most populous county in North Carolina with a 2020 census population of 541,299 ^13^. Examining core group composition in Guilford County provides insight on gonorrhea dynamics in regions of high gonorrhea rates outside of large metropolises.

## Methods

### Cohort

The CDC launched a multi-site program SURRG (Strengthening the U.S. Response to Resistant Gonorrhea) in 2016 in order to rapidly detect and respond to cases of drug-resistant gonorrhea^10^. Guilford County Department of Health and Human Services Division of public health was selected as one of the SURRG surveillance locations and consisted of a consortium of two STI clinics and 4 non-STI clinics conducting surveillance activities for resistant *N. gonorrhoeae*. The cohort consists of patients who presented to a Guilford County SURRG site and were tested for *N. gonorrhoeae* from January 2018 to December 2023.

### Data collection

Specific data are collected on individuals with specimens collected by SURRG including certain demographic, health, and encounter information, behavior risks, and longitudinal testing results. Demographic, health, and encounter information included date of encounter, clinic, patient’s age, sexual orientation, sex at birth, ethnicity, race, pregnancy status, previous gonorrhea infection, HIV status, and current symptoms. Patients were interviewed on intake to obtain reported sexual risk behaviors. Finally longitudinal *N. gonorrhoeae* diagnostic results were captured with cases being defined as positive laboratory confirmation either with a culture or a positive nucleic acid amplification test (NAAT) from a sample of any location (i.e. genital, extra-genital).

### Outcome definitions

Patients were determined to be in the core group if they had multiple known gonorrhea infections: those that tested positive for gonorrhea on multiple clinic visits (at least 21 days between positive events), and individuals that tested positive and indicated they had had a previous infection.^8,9^ Three primary subpopulations in the data are explored: teens (n = 3,170) are considered any patient that was 19 years or younger at their first visit; adult MSMs (n = 1,521) are any 20+ year old men who report having sex with men; adult heterosexuals (n= 14,009) are any 20+ year old individuals who report opposite sex partners only.

### Statistical analysis

All statistical analyses were performed using R (version 4.4.0). Summary statistics for each sub-population, as well as the overall population (included groups not represented in the explored sub-populations such as adults without known sexual partners and adult women who have sex with women) were calculated. These included means and standard deviations for continuous variables, and frequencies and percentages for categorical variables. To assess the likelihood of being included in the core group, odds ratios (ORs) were calculated using univariate logistic regression models using maximum likelihood. The ORs were adjusted for potential confounders, and confidence intervals (CI) were computed to determine the precision of the estimates.

Statistical significance was set at p<0.05 for all tests.

## Results

Our study included 25,520 SURRG patients from January 2018 to December 2023, of whom 1,718 (6.7%) were classified as part of the core group. Demographic details of the study population are presented in Table 1. Within the core group, men were disproportionately represented, constituting 60% of the core group, compared to 29% of the total patient population. The core group was composed of 18.5% teens, 51.2% heterosexual adults, 19.2% MSM adults, and 11% other adults.

**Table 1:**
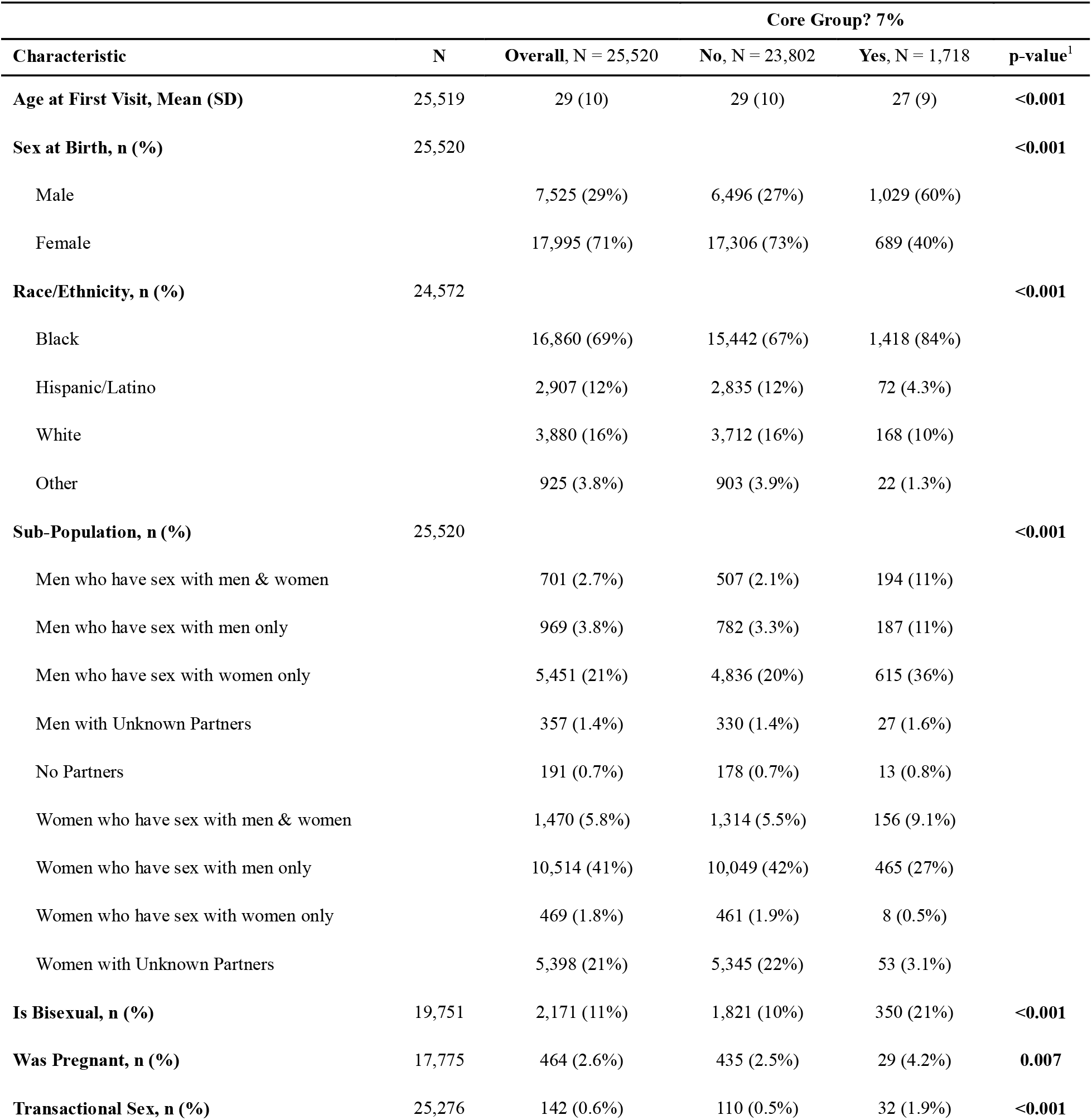
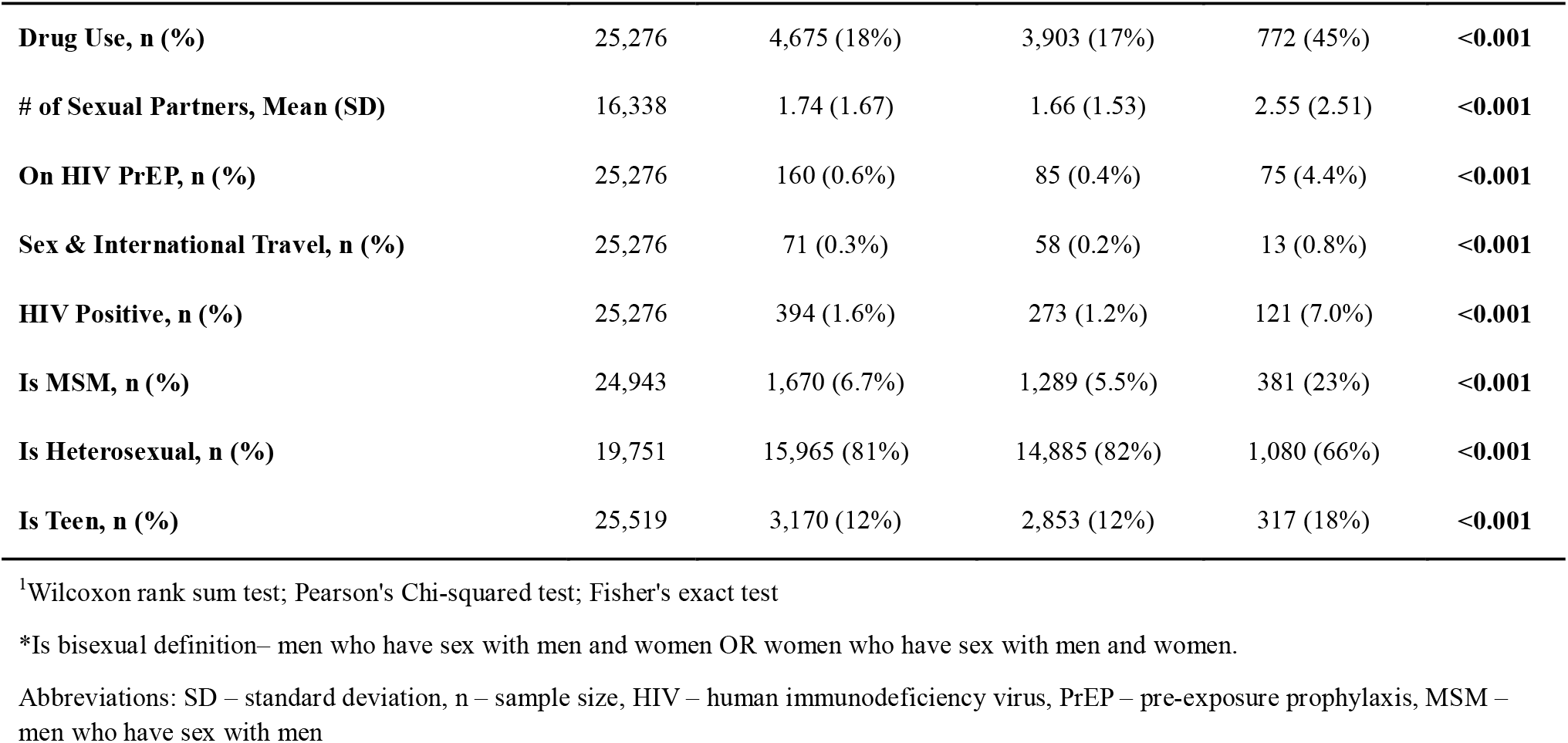
Summary table of SURRG cohort both in and out of the core group 2018-2023.

The core group represented 7% of the total patient population but accounted for 68% of all gonorrhea cases. Within subgroups, the teen core group comprised 10% of all teens and accounted for 66% of teen gonorrhea cases. The adult MSM core group represented 22% of all adult MSM patients and accounted for 79% of adult MSM gonorrhea cases. The adult heterosexual core group made up 6% of all adult heterosexuals and accounted for 67% of adult heterosexual gonorrhea cases. A graphical comparison of the distribution of patients and cases for each subpopulation is provided in Figure 1.

**Figure 1:**
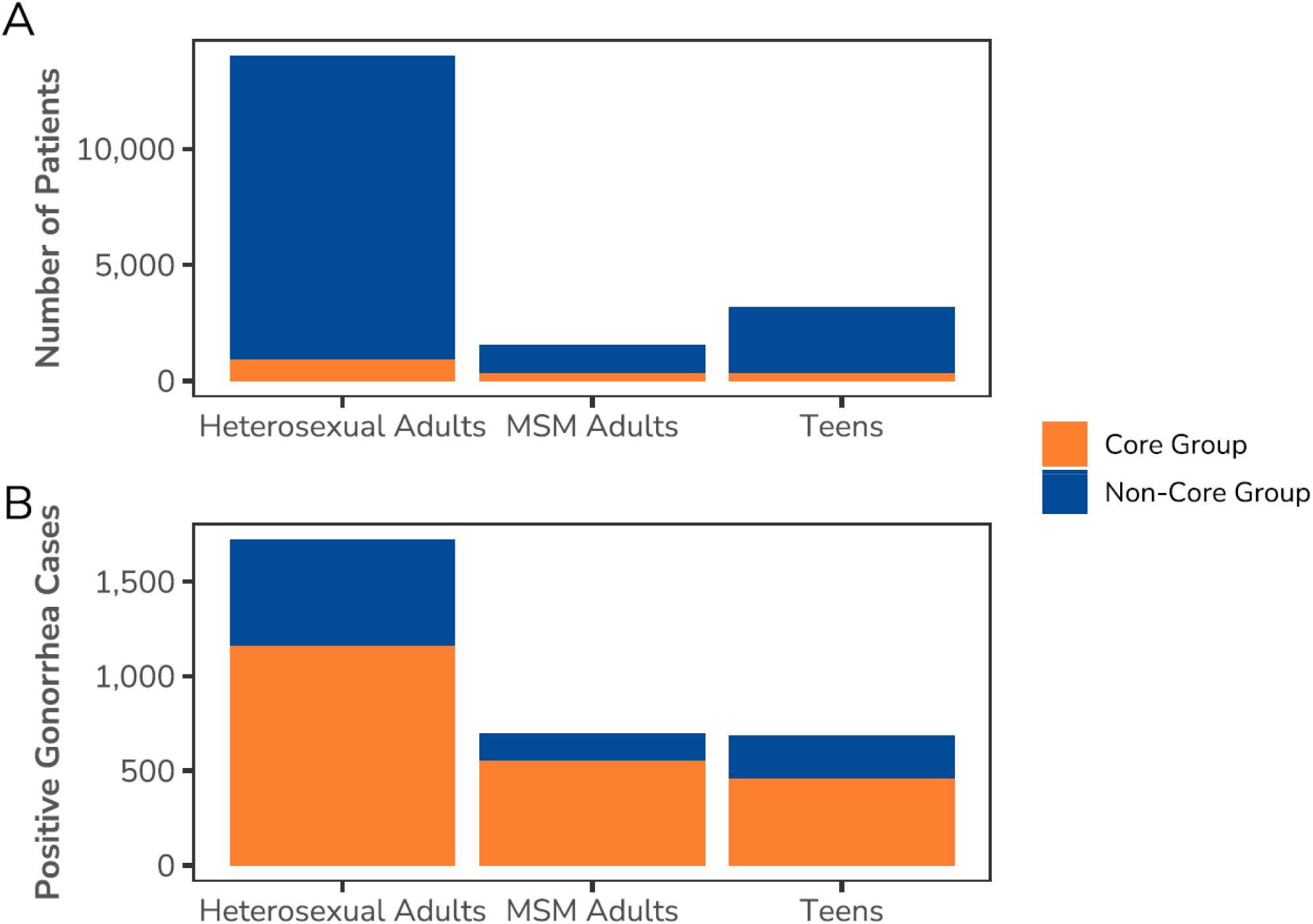
Distribution of patients and cases across subpopulations for the SURRG cohort collected from 2018-2023. Panel A shows the breakdown of core and non-core groups within the total SURRG population across the subgroups of interest: heterosexual adults, MSM adults, and teens. Panel B illustrates the distribution of core and non-core groups specifically for gonorrhea-positive patients within the SURRG cohort.

Full descriptive statistics for each subpopulation are available in the supplement (Supplement Tables 1-3). The distribution of sexual partners within each subgroup is shown in Figure 2. Notably, the MSM subgroup exhibited a heavier right tail, indicating that a larger proportion of MSM individuals reported a higher number of sexual partners compared to other subpopulations. Additional demographic and behavioral risk factors are summarized in Table 2, with odds ratios provided for both the general population and each subpopulation.

**Figure 2:**
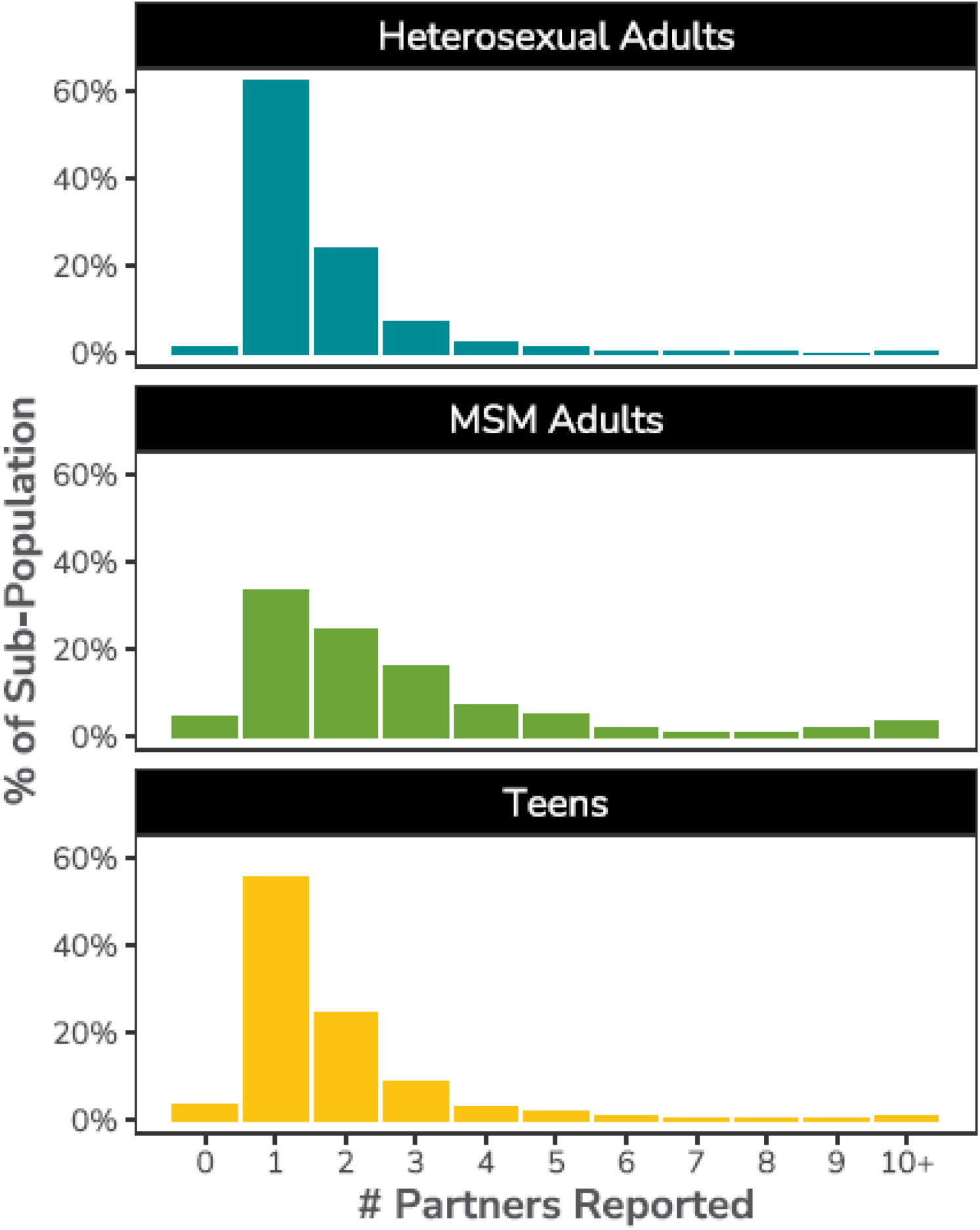
Distribution of sexual partners within each subgroup. The distribution of sexual partners for each subgroup is shown, with a focus on the differences across subpopulations. The top panel represents the heterosexual adult subgroup, the middle panel represents the MSM adult subgroup, and the bottom panel shows the teen subpopulation.

**Table 2:**
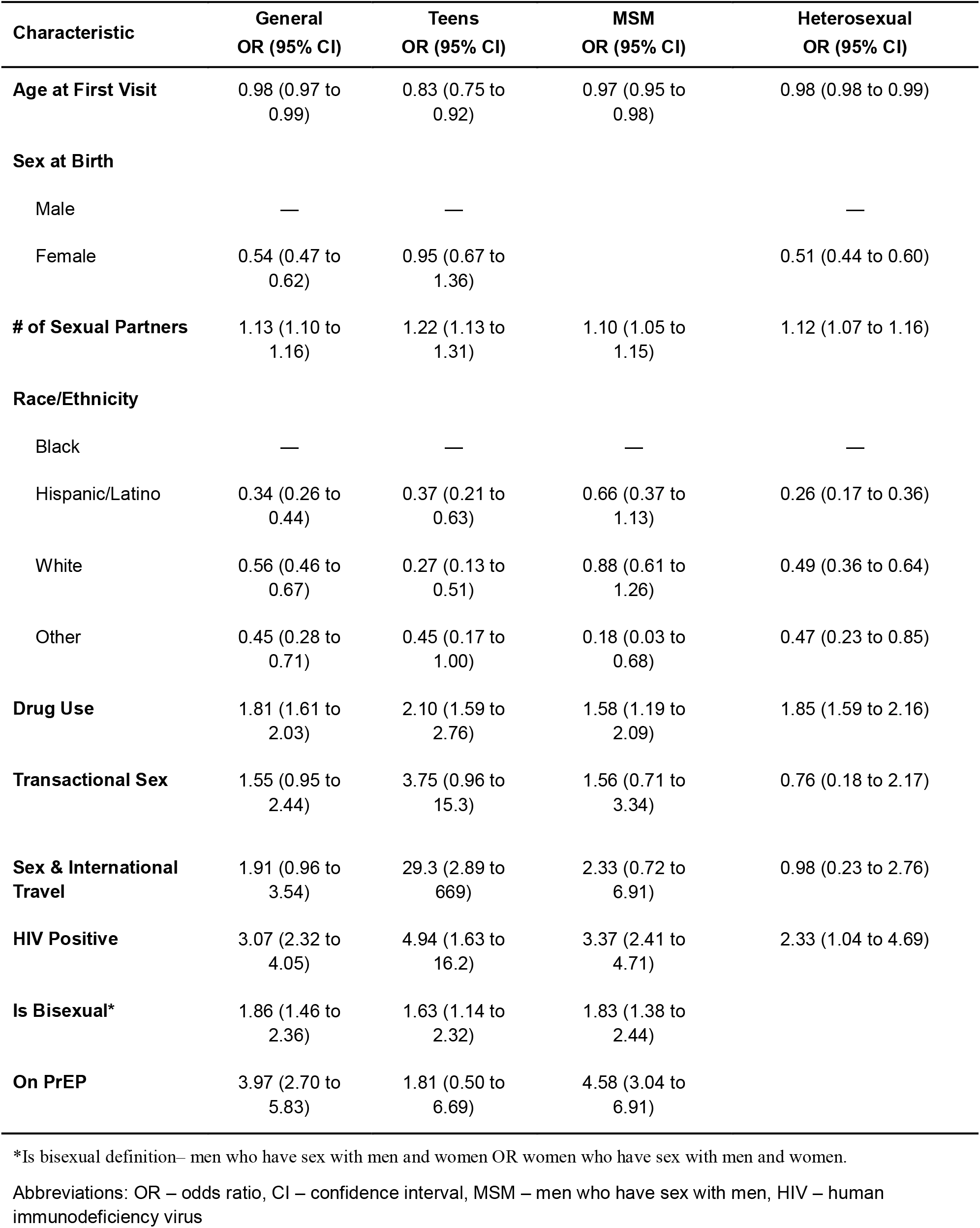
Regression table for different subgroups within the SURRG population.

| Characteristic | General<br>OR (95% CI) | Teens<br>OR (95% CI) | MSM<br>OR (95% CI) | Heterosexual<br>OR (95% CI) |
| --- | --- | --- | --- | --- |
| **Age at First Visit** | 0.98 (0.97 to 0.99) | 0.83 (0.75 to 0.92) | 0.97 (0.95 to 0.98) | 0.98 (0.98 to 0.99) |
| **Sex at Birth** |  |  |  |  |
| Male | — | — |  | — |
| Female | 0.54 (0.47 to 0.62) | 0.95 (0.67 to 1.36) |  | 0.51 (0.44 to 0.60) |
| **# of Sexual Partners** | 1.13 (1.10 to 1.16) | 1.22 (1.13 to 1.31) | 1.10 (1.05 to 1.15) | 1.12 (1.07 to 1.16) |
| **Race/Ethnicity** |  |  |  |  |
| Black | — | — | — | — |
| Hispanic/Latino | 0.34 (0.26 to 0.44) | 0.37 (0.21 to 0.63) | 0.66 (0.37 to 1.13) | 0.26 (0.17 to 0.36) |
| White | 0.56 (0.46 to 0.67) | 0.27 (0.13 to 0.51) | 0.88 (0.61 to 1.26) | 0.49 (0.36 to 0.64) |
| Other | 0.45 (0.28 to 0.71) | 0.45 (0.17 to 1.00) | 0.18 (0.03 to 0.68) | 0.47 (0.23 to 0.85) |
| **Drug Use** | 1.81 (1.61 to 2.03) | 2.10 (1.59 to 2.76) | 1.58 (1.19 to 2.09) | 1.85 (1.59 to 2.16) |
| **Transactional Sex** | 1.55 (0.95 to 2.44) | 3.75 (0.96 to 15.3) | 1.56 (0.71 to 3.34) | 0.76 (0.18 to 2.17) |
| **Sex & International Travel** | 1.91 (0.96 to 3.54) | 29.3 (2.89 to 669) | 2.33 (0.72 to 6.91) | 0.98 (0.23 to 2.76) |
| **HIV Positive** | 3.07 (2.32 to 4.05) | 4.94 (1.63 to 16.2) | 3.37 (2.41 to 4.71) | 2.33 (1.04 to 4.69) |
| **Is Bisexual\*** | 1.86 (1.46 to 2.36) | 1.63 (1.14 to 2.32) | 1.83 (1.38 to 2.44) |  |
| **On PrEP** | 3.97 (2.70 to 5.83) | 1.81 (0.50 to 6.69) | 4.58 (3.04 to 6.91) |  |

\*Is bisexual definition— men who have sex with men and women OR women who have sex with men and women.
Abbreviations: OR – odds ratio, CI – confidence interval, MSM – men who have sex with men, HIV – human immunodeficiency virus

## Discussion

Gonorrhea dynamics are known to vary significantly by geography and population^7,14–19^. Our study, conducted in Guilford County, North Carolina, provides valuable insight into these regional differences. In North Carolina, the rate of gonorrhea was similar in 2023 to 2022; there were 26,382 gonorrhea cases in 2023 (rate of 246.6 cases per 100,000 population)^20^. North Carolina is located in the Southeast United States, a region that consistently reports the highest gonorrhea rates^11^. Guilford County, which is home to approximately 541,299 people, includes the major cities of Greensboro and High Point, both of which are within the Piedmont Triad metropolitan area^13^. The region is home to persons from varying backgrounds, concentrated risks for poor outcomes, and health care deserts. diverse demographic composition, with a significant proportion of Black residents, particularly in the historically segregated Eastern Greensboro area, where socioeconomic disparities are marked by lower life expectancy, higher poverty rates, and limited access to healthcare^13^. These factors may exacerbate the spread of sexually transmitted infections like gonorrhea, particularly in neighborhoods that face concentrated disadvantage. Our findings in this context underscore the importance of considering regional and demographic factors when addressing gonorrhea prevention and treatment, particularly in areas like Guilford County, where higher rates of infection may be linked to both geographic and social determinants of health. Furthermore, our study offers important insights into the teen core group composition and associated risk factors, which are important given the growth of STIs in younger age groups^11,20^.

While the study provides valuable insights into gonorrhea transmission dynamics in Guilford County, several limitations in the methods should be considered. First, the cohort is limited to individuals who presented at participating SURRG clinics, which may not fully represent the broader population of those at risk for gonorrhea in the community, especially those who do not seek care or who access healthcare outside of these sites. While behavioral risk data were collected through provider conducted interviews, self-reported data may be subject to recall bias or social desirability bias, potentially leading to underreporting or misclassification of certain risk behaviors. Finally, while providing insights into gonorrhea dynamics in Guilford County, these findings are not generalizable to other populations.

Numerous studies have identified higher rates of gonorrhea infections and an increased risk of being in the gonorrhea core among Black populations^14,15,17,19,21^. Work by Thomas and Gaffield examining counties in the US southeast from 1986-1995 with high endemic levels of gonorrhea showed that the association of larger proportions of Black individuals in a population and high gonorrhea rates was mediated by social structure factors such as racial residential isolation and black-white income dualism^22^. Our calculated odds ratios align with these findings across the general population, teens, and heterosexual adults, reaffirming the disproportionate burden faced by Black individuals in the southeastern United States. However, in our study, among MSM the odds ratios associated with being White or Hispanic/Latino was not significantly protective compared to the Black reference group. This suggests that the MSM subcommunity in Guilford County may exhibit distinct epidemiological patterns compared to other regions, warranting further investigation into the unique transmission dynamics and risk factors within this group.

Higher sexually transmitted infection rates as well as emerging antimicrobial resistant trends are associated with MSM populations.^8,23^ These increasing rates are associated with demographic changes, sexual risk behaviors, and the sociocultural environment.^24^ Our findings in Guilford County fit within these broader trends. In addition to being a major component of the core group, the distribution for the MSM subgroup reported number of sexual partners had a heavier right tail compared to the other subpopulations, indicating a greater proportion of MSM reporting a higher number of sexual partners. This suggests that within the MSM subgroup, there is a subset of individuals engaging in higher levels of sexual activity, which may contribute to the observed elevated risk of gonorrhea transmission. These trends underscore the need for targeted interventions aimed at reducing sexual risk in this group, particularly considering the intersection of high-risk sexual behavior and the elevated odds ratio for gonorrhea in this subgroup.

In the context of gonorrhea transmission dynamics, adolescents have often been an understudied population, despite their significant role in the transmission of sexually transmitted infections. A study by Kaufman et al. looking at gonorrhea cases nationally using national data from Quest Diagnostics found a decrease prevalence in gonorrhea positivity in teens aged 12-17 years, but an increasing trend for the 18-24 years age group^25^. Despite the overall decline in gonorrhea positivity in this younger cohort nationally, our findings suggest that teenagers remain an important demographic in the local core group, particularly in high-transmission settings like Guilford County. These results are consistent with trends across North Carolina; rates in the 15-19 year old age group are increasing for the state.^20^ Women in this age group with gonorrhea infections are at much higher risk of pelvic inflammatory diseases and adverse pregnancy outcomes.^26^

Our study results underscore the importance of teens as a critical sub-population within gonorrhea core groups, particularly in the southeastern United States. These findings highlight the need for more focused surveillance and research on this age group to better understand the patterns and determinants of transmission. Our analysis identifies key factors associated with inclusion in the gonorrhea core (or having repeated gonorrhea infections), which could inform targeted prevention and intervention strategies aimed at reducing recurrence in this vulnerable demographic. We assessed the odds of being included in the gonorrhea core group among individuals who reported engaging in sex during international travel. Although the overall odds ratio was not statistically significant for the overall study cohort, the MSM adult subgroup, or the heterosexual adult subgroup, a strikingly high odds ratio of 29.3 was found in the teen subgroup. This suggests that international travel-related sexual activity may represent a significant risk factor for gonorrhea transmission, particularly in adolescents, who may have higher vulnerability due to factors such as sexual risk behavior and limited access to preventive healthcare. Further research is needed to explore the underlying factors contributing to this increased risk in teens and to develop targeted interventions for this high-risk group.

To address the ongoing challenge of gonorrhea transmission, interventions targeting the core group—individuals at highest risk of repeated infections—are crucial. Focused approaches that prioritize the MSM subgroup, adolescents, and populations experiencing social and structural disadvantage can play a pivotal role in curbing the spread of gonorrhea. Community-based outreach, educational programs, and increased access to preventive healthcare, including regular screening (including multimodal testing such as at-home or self-collection) and treatment, are essential in reducing transmission rates in these high-risk groups. Integrating strategies such as doxycycline postexposure prophylaxis and promoting safer sexual practices within the core group could complement efforts to mitigate the burden of gonorrhea. Doxycycline postexposure prophylaxis (PEP), show promise in reducing bacterial STI transmission, particularly among populations at higher risk, including MSM^27^. Studies demonstrate that doxyPEP significantly decreases the incidence of syphilis, chlamydia, and gonorrhea when administered within 72 hours of exposure^27–29^ However, balancing the effectiveness of these interventions with the potential risk of developing antimicrobial resistance is vital. Studies have found that targeting core groups with antimicrobial therapeutics, though key to reducing transmission, can drive antimicrobial resistance^8,30^. Future research should explore optimal intervention strategies that can reduce transmission while minimizing resistance, ensuring sustainable public health outcomes.

## Data Availability

Individual-level data collected as part of Neisseria gonorrhea surveillance activity is not publicly available. All data relevant to this study were presented in this article.

## Acknowledgements

We would like to gratefully acknowledge all members of the SURRG-Guilford County team, whose collective expertise, collaboration, and dedication were instrumental in the success of this project. In particular, we acknowledge the contributions of Guilford County Public Health staff including Dianne Jackson, Chanelle McChristian, Latonya Pender, Brandy Svedek and Parrish Webster-Sloan. At the North Carolina Department of Health and Human Services, William Glover, Erika Samoff, and Victoria Mobley supported the project. Finally, at Wake Forest University School of Medicine, Kara Libby, Jennifer Nall, Elizabeth Palavecino, and Kim Reeves contributed to the success of the project.

## Supplement

**Supplement Table 1:** Teen subgroup summary table

**Supplement Table 2:** MSM adult subgroup summary table

**Supplement Table 3:** Heterosexual adult subgroup summary table

## References

1. Hadeler KP, Castillo-Chavez C. A core group model for disease transmission. Math Biosci. 1995;b128(1-2):41–55. doi:10.1016/0025-5564(94)00066-9

2. Yorke JA, Hethcote HW, Nold A. Dynamics and control of the transmission of gonorrhea. Sex Transm Dis. 1978;5(2):51–56. doi:10.1097/00007435-197804000-00003

3. Endo A, Murayama H, Abbott S, et al. Heavy-tailed sexual contact networks and monkeypox epidemiology in the global outbreak, 2022. Science. 2022;378(6615):90–94. doi:10.1126/science.add4507

4. Anderson RM, Swinton J, Garnett GP. Potential impact of low efficacy HIV-1 vaccines in populations with high rates of infection. Proc Biol Sci. 1995;261(1361):147–151. doi:10.1098/rspb.1995.0129

5. Allan-Blitz LT, Klausner JD. Prevalence of mpox immunity among the core group and its potential to prevent future large-scale outbreaks. Lancet Microbe. 2024;5(12). doi:10.1016/j.lanmic.2024.100957

6. Palk L, Blower S. Geographic variation in sexual behavior can explain geospatial heterogeneity in the severity of the HIV epidemic in Malawi. BMC Med. 2018;16(1):22. doi:10.1186/s12916-018-1006-x

7. Rothenberg RB. The geography of gonorrhea. Empirical demonstration of core group transmission. Am J Epidemiol. 1983;117(6):688–694. doi:10.1093/oxfordjournals.aje.a113602

8. Lewis DA. The role of core groups in the emergence and dissemination of antimicrobial-resistant N gonorrhoeae. Sex Transm Infect. 2013;89 Suppl 4:iv47–51. doi:10.1136/sextrans-2013-051020

9. Thomas JC, Tucker MJ. The development and use of the concept of a sexually transmitted disease core. J Infect Dis. 1996;174 Suppl 2:S134–143. doi:10.1093/infdis/174.supplement_2.s134

10. Schlanger K, Learner ER, Pham CD, et al. Strengthening the US Response to Resistant Gonorrhea: An Overview of a Multisite Program to Enhance Local Response Capacity for Antibiotic-Resistant Neisseria gonorrhoeae. Sex Transm Dis. 2021;48(12S Suppl 2):S97–S103. doi:10.1097/OLQ.0000000000001545

11. Centers for Disease Control and Prevention. Sexually Transmitted Infections Surveillance 2023. US Department of Health and Human Services; 2024. https://www.cdc.gov/sti-statistics/annual/index.html

12. Martín-Sánchez M, Ong JJ, Fairley CK, et al. Clinical presentation of asymptomatic and symptomatic heterosexual men who tested positive for urethral gonorrhoea at a sexual health clinic in Melbourne, Australia. BMC Infect Dis. 2020;20(1):486. doi:10.1186/s12879-020-05197-y

13. U.S. Census Bureau. QuickFacts: Guilford County, North Carolina. U.S. Census Bureau; 2024. Accessed March 12, 2025. https://www.census.gov/quickfacts/fact/table/guilfordcountynorthcarolina/POP010220#POP0 10220

14. Potterat JJ, Rothenberg RB, Woodhouse DE, Muth JB, Pratts CI, Fogle JS. Gonorrhea as a Social Disease. Sex Transm Dis. 1985;12(1):25–32.

15. Rice RJ, Roberts PL, Handsfield HH, Holmes KK. Sociodemographic distribution of gonorrhea incidence: implications for prevention and behavioral research. Am J Public Health. 1991;81(10):1252–1258. doi:10.2105/AJPH.81.10.1252

16. Du P, McNutt LA, O’Campo P, Coles FB. Changes in Community Socioeconomic Status and Racial Distribution Associated With Gonorrhea Rates: An Analysis at the Community Level. Sex Transm Dis. 2009;36(7):430. doi:10.1097/OLQ.0b013e31819b8c2f

17. Monteiro EF, Lacey CJN, Merrick D. The interrelation of demographic and geospatial risk factors between four common sexually transmitted diseases. Sex Transm Infect. 2005;81(1):41–46. doi:10.1136/sti.2004.009431

18. Becker KM, Glass GE, Brathwaite W, Zenilman JM. Geographic Epidemiology of Gonorrhea in Baltimore, Maryland, Using a Geographic Information System. Am J Epidemiol. 1998;147(7):709–716. doi:10.1093/oxfordjournals.aje.a009513

19. Hamers FF, Peterman TA, Zaidi AA, Ransom RL, Wroten JE, Witte JJ. Syphilis and gonorrhea in Miami: similar clustering, different trends. Am J Public Health. 1995;85(8 Pt 1):1104–1108.

20. North Carolina HIV/STD/Hepatitis Surveillance Unit. 2023 North Carolina STD Surveillance Report. North Carolina Department of Health and Human Services, Division of Public Health, Communicable Disease Branch.; 2024. Accessed April 7, 2025. https://epi.dph.ncdhhs.gov/cd/stds/figures/2023-NC-STD-SurveillanceReportSummaryFinal.pdf

21. Owusu-Edusei K, Chesson HW, Leichliter JS, Kent CK, Aral SO. The Association Between Racial Disparity in Income and Reported Sexually Transmitted Infections. Am J Public Health. 2013;103(5):910–916. doi:10.2105/AJPH.2012.301015

22. Thomas JC, Gaffield ME. Social Structure, Race, and Gonorrhea Rates in the Southeastern United States. Ethn Dis. 2003;13(3):362–368.

23. Unemo M, Bradshaw CS, Hocking JS, et al. Sexually transmitted infections: challenges ahead. Lancet Infect Dis. 2017;17(8):e235–e279. doi:10.1016/S1473-3099(17)30310-9

24. Fenton KA, Imrie J. Increasing Rates of Sexually Transmitted Diseases in Homosexual Men in Western Europe and the United States: Why? Infect Dis Clin. 2005;19(2):311–331. doi:10.1016/j.idc.2005.04.004

25. Kaufman HW, Gift TL, Kreisel K, Niles JK, Alagia DP. Chlamydia and Gonorrhea: Shifting Age-Based Positivity Among Young Females, 2010–2017. Am J Prev Med. 2020;59(5):697–703. doi:10.1016/j.amepre.2020.05.023

26. Vallely LM, Egli-Gany D, Wand H, et al. Adverse pregnancy and neonatal outcomes associated with Neisseria gonorrhoeae: systematic review and meta-analysis. Sex Transm Infect. 2021;97(2):104–111. doi:10.1136/sextrans-2020-054653

27. Bachmann LH. CDC Clinical Guidelines on the Use of Doxycycline Postexposure Prophylaxis for Bacterial Sexually Transmitted Infection Prevention, United States, 2024. MMWR Recomm Rep. 2024;73. doi:10.15585/mmwr.rr7302a1

28. Luetkemeyer AF, Donnell D, Dombrowski JC, et al. Postexposure Doxycycline to Prevent Bacterial Sexually Transmitted Infections. N Engl J Med. 2023;388(14):1296–1306. doi:10.1056/NEJMoa2211934

29. Molina JM, Charreau I, Chidiac C, et al. Post-exposure prophylaxis with doxycycline to prevent sexually transmitted infections in men who have sex with men: an open-label randomised substudy of the ANRS IPERGAY trial. Lancet Infect Dis. 2018;18(3):308–317. doi:10.1016/S1473-3099(17)30725-9

30. Chan CH, McCabe CJ, Fisman DN. Core groups, antimicrobial resistance and rebound in gonorrhoea in North America. Sex Transm Infect. 2012;88(3):200–204. doi:10.1136/sextrans-2011-050049

