## Supplement for "Core group dynamics of gonorrhea infection in a North Carolina county from 2018 to 2023"

**Supplement Table 1:** Teen subgroup summary table

|  | | | **Core Group? 10%** | |  |
| --- | --- | --- | --- | --- | --- |
| **Characteristic** | **N** | **Overall**, N = 3,170 | **No**, N = 2,853 | **Yes**, N = 317 | **p-value**^1^ |
| **Age at First Visit, n (%)** | 3,170 |  |  |  |  |
| 11 |  | 2 (<0.1%) | 2 (<0.1%) | 0 (0%) |  |
| 12 |  | 2 (<0.1%) | 2 (<0.1%) | 0 (0%) |  |
| 13 |  | 11 (0.3%) | 11 (0.4%) | 0 (0%) |  |
| 14 |  | 33 (1.0%) | 29 (1.0%) | 4 (1.3%) |  |
| 15 |  | 144 (4.5%) | 129 (4.5%) | 15 (4.7%) |  |
| 16 |  | 261 (8.2%) | 229 (8.0%) | 32 (10%) |  |
| 17 |  | 488 (15%) | 425 (15%) | 63 (20%) |  |
| 18 |  | 941 (30%) | 855 (30%) | 86 (27%) |  |
| 19 |  | 1,288 (41%) | 1,171 (41%) | 117 (37%) |  |
| **Sex at Birth, n (%)** | 3,170 |  |  |  | **<0.001** |
| Male |  | 582 (18%) | 469 (16%) | 113 (36%) |  |
| Female |  | 2,588 (82%) | 2,384 (84%) | 204 (64%) |  |
| **Race/Ethnicity, n (%)** | 3,043 |  |  |  | **<0.001** |
| Black |  | 2,171 (71%) | 1,902 (70%) | 269 (87%) |  |
| Hispanic/Latino |  | 335 (11%) | 317 (12%) | 18 (5.8%) |  |
| White |  | 391 (13%) | 377 (14%) | 14 (4.5%) |  |
| Other |  | 146 (4.8%) | 139 (5.1%) | 7 (2.3%) |  |
| **Sub-Population, n (%)** | 3,170 |  |  |  |  |
| Men who have sex with men & women |  | 66 (2.1%) | 39 (1.4%) | 27 (8.5%) |  |
| Men who have sex with men only |  | 83 (2.6%) | 60 (2.1%) | 23 (7.3%) |  |
| Men who have sex with women only |  | 387 (12%) | 326 (11%) | 61 (19%) |  |
| Men with Unknown Partners |  | 41 (1.3%) | 40 (1.4%) | 1 (0.3%) |  |
| No Partners |  | 40 (1.3%) | 35 (1.2%) | 5 (1.6%) |  |
| Women who have sex with men & women |  | 242 (7.6%) | 191 (6.7%) | 51 (16%) |  |
| Women who have sex with men only |  | 1,569 (49%) | 1,430 (50%) | 139 (44%) |  |
| Women who have sex with women only |  | 51 (1.6%) | 48 (1.7%) | 3 (0.9%) |  |
| Women with Unknown Partners |  | 691 (22%) | 684 (24%) | 7 (2.2%) |  |
| **Is Bisexual, n (%)** | 2,433 | 308 (13%) | 230 (11%) | 78 (25%) | **<0.001** |
| **Was Pregnant, n (%)** | 2,568 | 61 (2.4%) | 48 (2.0%) | 13 (6.4%) | **<0.001** |
| **Transactional Sex, n (%)** | 3,149 | 12 (0.4%) | 6 (0.2%) | 6 (1.9%) | **<0.001** |
| **Drug Use, n (%)** | 3,149 | 786 (25%) | 608 (21%) | 178 (56%) | **<0.001** |
| **# of Sexual Partners, Mean (SD)** | 2,046 | 1.78 (1.70) | 1.64 (1.38) | 2.58 (2.83) | **<0.001** |
| **On PrEP, n (%)** | 3,149 | 15 (0.5%) | 7 (0.2%) | 8 (2.5%) | **<0.001** |
| **Sex & International Travel, n (%)** | 3,149 | 4 (0.1%) | 1 (<0.1%) | 3 (0.9%) | **0.004** |
| **HIV Positive, n (%)** | 3,149 | 18 (0.6%) | 8 (0.3%) | 10 (3.2%) | **<0.001** |
| ^1^Pearson's Chi-squared test; Fisher's exact test; Wilcoxon rank sum test | | | | | |

**Supplement Table 2:** MSM adult subgroup summary table

|  | | | **Core Group? 22%** | |  |
| --- | --- | --- | --- | --- | --- |
| **Characteristic** | **N** | **Overall**, N = 1,521 | **No**, N = 1,190 | **Yes**, N = 331 | **p-value**^1^ |
| **Age at First Visit, Mean (SD)** | 1,521 | 31 (11) | 32 (11) | 29 (9) | **0.002** |
| **Race/Ethnicity, n (%)** | 1,463 |  |  |  | **<0.001** |
| Black |  | 940 (64%) | 705 (62%) | 235 (73%) |  |
| Hispanic/Latino |  | 138 (9.4%) | 119 (10%) | 19 (5.9%) |  |
| White |  | 352 (24%) | 284 (25%) | 68 (21%) |  |
| Other |  | 33 (2.3%) | 31 (2.7%) | 2 (0.6%) |  |
| **Sub-Population, n (%)** | 1,521 |  |  |  | **<0.001** |
| Men who have sex with men & women |  | 635 (42%) | 468 (39%) | 167 (50%) |  |
| Men who have sex with men only |  | 886 (58%) | 722 (61%) | 164 (50%) |  |
| **Is Bisexual, n (%)** | 1,521 | 635 (42%) | 468 (39%) | 167 (50%) | **<0.001** |
| **Transactional Sex, n (%)** | 1,521 | 36 (2.4%) | 21 (1.8%) | 15 (4.5%) | **0.003** |
| **Drug Use, n (%)** | 1,521 | 522 (34%) | 362 (30%) | 160 (48%) | **<0.001** |
| **# of Sexual Partners, Mean (SD)** | 1,454 | 2.78 (3.19) | 2.52 (2.93) | 3.70 (3.84) | **<0.001** |
| **On PrEP, n (%)** | 1,521 | 138 (9.1%) | 71 (6.0%) | 67 (20%) | **<0.001** |
| **Sex & International Travel, n (%)** | 1,521 | 18 (1.2%) | 12 (1.0%) | 6 (1.8%) | 0.25 |
| **HIV Positive, n (%)** | 1,521 | 272 (18%) | 176 (15%) | 96 (29%) | **<0.001** |
| ^1^Wilcoxon rank sum test; Pearson's Chi-squared test; Fisher's exact test | | | | | |

**Supplement Table 3:** Heterosexual adult subgroup summary table

|  | | | **Core Group? 6%** | |  |
| --- | --- | --- | --- | --- | --- |
| **Characteristic** | **N** | **Overall**, N = 14,009 | **No**, N = 13,129 | **Yes**, N = 880 | **p-value**^1^ |
| **Age at First Visit, Mean (SD)** | 14,009 | 31 (9) | 31 (9) | 29 (9) | **<0.001** |
| **Sex at Birth, n (%)** | 14,009 |  |  |  | **<0.001** |
| Male |  | 5,064 (36%) | 4,510 (34%) | 554 (63%) |  |
| Female |  | 8,945 (64%) | 8,619 (66%) | 326 (37%) |  |
| **Race/Ethnicity, n (%)** | 13,715 |  |  |  | **<0.001** |
| Black |  | 9,533 (70%) | 8,764 (68%) | 769 (89%) |  |
| Hispanic/Latino |  | 1,909 (14%) | 1,879 (15%) | 30 (3.5%) |  |
| White |  | 1,879 (14%) | 1,822 (14%) | 57 (6.6%) |  |
| Other |  | 394 (2.9%) | 384 (3.0%) | 10 (1.2%) |  |
| **Sub-Population, n (%)** | 14,009 |  |  |  | **<0.001** |
| Men who have sex with women only |  | 5,064 (36%) | 4,510 (34%) | 554 (63%) |  |
| Women who have sex with men only |  | 8,945 (64%) | 8,619 (66%) | 326 (37%) |  |
| **Was Pregnant, n (%)** | 8,945 | 359 (4.0%) | 350 (4.1%) | 9 (2.8%) | 0.24 |
| **Transactional Sex, n (%)** | 14,009 | 51 (0.4%) | 48 (0.4%) | 3 (0.3%) | >0.99 |
| **Drug Use, n (%)** | 14,009 | 2,857 (20%) | 2,479 (19%) | 378 (43%) | **<0.001** |
| **# of Sexual Partners, Mean (SD)** | 11,442 | 1.61 (1.29) | 1.57 (1.27) | 2.12 (1.46) | **<0.001** |
| **On PrEP, n (%)** | 14,009 | 0 (0%) | 0 (0%) | 0 (0%) |  |
| **Sex & International Travel, n (%)** | 14,009 | 41 (0.3%) | 38 (0.3%) | 3 (0.3%) | 0.74 |
| **HIV Positive, n (%)** | 14,009 | 67 (0.5%) | 57 (0.4%) | 10 (1.1%) | **0.009** |
| ^1^Wilcoxon rank sum test; Pearson's Chi-squared test; Fisher's exact test | | | | | |
